# The Impact of Violence on Labour Force Participation and Income in Canada: A Cross-sectional Study with Linked Survey and Tax Data

**DOI:** 10.64898/2025.12.12.25342170

**Authors:** Gabriel John Dusing, Beverley M. Essue, Sujata Mishra, Nicholas Metheny, Candace Milinkovic, Felicia M. Knaul, Nata Duvvury

**Affiliations:** School of Health Policy and Management, York University, 4700 Keele Street, Toronto, ON M3J 1P3 Canada; Dalla Lana School of Public Health, Institute of Health Policy, Management and Evaluation, University of Toronto, 155 College Street, Toronto, ON M5T 3M6, Canada; Nell Hodgson Woodruff School of Nursing, Emory University, 1520 Clifton Road NE, Atlanta, GA 30322, United States of America; The University of California, Los Angeles (UCLA), David Geffen School of Medicine, 885 Tiverton Drive Los Angeles, California 90095, United States of America; School of Medicine and Health Sciences and Faculty of Excellence, TEC de Monterrey. Eugenio Garza Sada 2501, Tecnológico, 64700, Monterrey, Nuevo León, México; Tómatelo a Pecho A.C. Explanada 115, Lomas de Chapultepec sección I, 11000, Mexico City, Mexico; School of Political Science & Sociology, University of Galway, University Road, Galway, Ireland H91 TK33

## Abstract

**Background:** Violence across the life course is a persistent global problem with well-documented health and social consequences. Less is known about its relationship with labour market outcomes in high-income countries with strong social protections, such as Canada. This study examines whether lifetime exposure to physical or sexual violence is associated with labour force participation (LFP), reasons for economic inactivity, sectoral and occupational sorting, and income.

**Methods:** We analyzed data from the 2018 Canadian Survey on Safety in Public and Private Spaces (SSPPS), a nationally representative cross-sectional survey linked to 2018 administrative tax records. The analytic sample included working-age adults (18–64) with complete data on violence exposure and labour market outcomes. Lifetime violence exposure captured childhood abuse, adulthood non-partner violence, and intimate partner violence. Outcomes included past-year LFP, part- versus full-time work, employment sector and occupation, and annual personal income. We described labour market patterns by gender and exposure and used inverse probability weighted regression adjustment (IPWRA) to estimate average treatment effects (ATEs) on economic inactivity, using unexposed men as the reference group.

**Results:** Nearly 62 percent of respondents reported lifetime violence exposure (64.5 percent of women, 59.1 percent of men). Past-year labour force participation was high (85.9 percent overall) and showed minimal differences by exposure status: 82.0 percent of exposed women versus 80.3 percent of unexposed women, and 90.7 percent of exposed men versus 90.1 percent of unexposed men. IPWRA models indicated that, relative to unexposed men, exposed women had small but statistically significant increases in the probability of health-related inactivity (ATE: 0.009; 95%CI: 0.000-0.017) and early retirement (ATE: 0.015; 95%CI: 0.000 to 0.031), whereas ATEs for exposed men were small and non-significant across all outcomes. Sectoral and occupational distributions differed chiefly by gender; within-gender differences by exposure were limited. Income patterns were inconsistent by exposure status. For example, among women with secondary education or less, exposed women earned markedly less than unexposed women ($34,604 vs. $39,913), while differences among men were smaller and uniformly negative (exposed $74,981 vs. unexposed $75,208).

**Conclusions:** In Canada’s welfare-state context, lifetime violence exposure shows limited association with labour force participation but may influence specific pathways into inactivity and sectoral sorting. Longitudinal analyses are needed to clarify longer-term economic impacts.

## Introduction

Physical and sexual violence are persistent and pervasive issues worldwide (Cagney et al., 2025; Spencer et al., 2023). While men and boys are also affected, women and girls disproportionately experience these forms of violence across the life course (Heise et al., 2002; World Health Organization, 2021). Global estimates suggest that 16-25% of girls and 9-23% of boys experience violence in childhood (Cagney et al., 2025), and in adulthood, approximately 23-31% of women globally experience violence at least once in their lifetime (World Health Organization, 2021). In this study, we focus on physical and sexual violence, whether perpetrated by an intimate partner or non-partner, hereafter referred to as “violence,” while recognizing that the full spectrum of violence extends beyond these forms.

Violence has been linked to diminished physical and mental health (Spencer et al., 2023; Stein et al., 2025), poorer educational attainment (Assini-Meytin et al., 2022; Boden et al., 2007; Mitchell et al., 2021), and reduced long-term earnings potential (Assini-Meytin et al., 2022). Violence in adulthood, including in the context of intimate partner violence (IPV), can result in reduced labour force participation (LFP) and income (MacGregor et al., 2021; Sabia et al., 2013). Violence is thought to influence LFP through both direct and indirect pathways. Violence often has direct physical and mental health consequences, which can lead to long-term morbidities that impede participation in the workforce. Survivors often experience chronic pain, disability, post-traumatic stress disorder (PTSD), depression, and anxiety (Bosch et al., 2017; D’arcy-Bewick et al., 2022; Spencer et al., 2023), all of which can limit their ability to maintain stable employment. Perpetrators of violence may also restrict their partner’s autonomy by preventing them from working, controlling earnings, or sabotaging employment opportunities (Sabri et al., 2023; Stark & Hester, 2019; Steinert et al., 2023).

Violence can affect LFP indirectly by disrupting educational attainment, which in turn limits access to stable, well-paying employment. Research has shown that individuals exposed to violence over the life course, particularly childhood abuse, are more likely to experience poor educational outcomes, including lower graduation rates and reduced likelihood of higher education attainment (Boden et al., 2007; Fry et al., 2018; Geppert et al., 2024; Klencakova et al., 2023), which negatively affect long-term LFP and economic prospects. On the other hand, some survivors may increase their participation as a means of achieving financial independence after leaving a violent relationship, while others may be forced into economic self-sufficiency after experiencing violence in childhood.

Population-based studies suggest exposure to violence shapes labour market outcomes in complex, context-dependent ways. For example, research from the United States (US) (Henkhaus, 2022) and sub-Saharan Africa (Ouedraogo & Stenzel, 2021) links violence to poor economic outcomes, including lower earnings and reduced economic activity. In contrast, a population-based study from Turkey found that women had higher likelihood of LFP (Gedikli et al., 2023). Furthermore, the economic consequences for survivors extend beyond participation to include impacts on productivity. A study in Peru, for example, found that employed survivors had significantly higher odds of workplace absenteeism and presenteeism (Asencios-Gonzalez et al., 2024). These combined impacts on both participation and productivity can deepen personal financial insecurity and, when aggregated, limit overall economic growth (Duvvury et al., 2023; Raghavendra et al., 2017). However, the generalizability of these findings to a high-income country with a robust welfare state and distinct labour market structures, such as Canada, remains unclear.

The Canadian context is distinct from the aforementioned for several reasons. First, although the US and Canada are both characterized as liberal welfare regimes (Myles, 1998) with similar levels of social spending as a percentage of gross domestic product (Béland et al., 2021), they have some key differences. Canadian social policy relies on universal benefits and services, whereas the US emphasizes a mix of means-tested social assistance and insurance (Béland et al., 2021; Béland & Waddan, 2017). Second, Canadian women’s labour force participation has steadily increased since the 1990s, with fewer exits after childbirth (Moyser, 2019). Moreover, Canadian women hold higher rates of post-secondary degrees than men (Moyser, 2019) and are represented in diverse occupational categories (Pelletier et al., 2019). At the same time, women in Canada face persistent challenges related to violence despite prevention and policy efforts across federal (Women and Gender Equality Canada, 2024a, 2024b) and provincial (Gouvernement du Québec, 2024; Government of Alberta, 2025; Government of Nova Scotia, 2023) jurisdictions. Approximately 44% of Canadian women report experiencing some form of violence at some point in their lives (Cotter, 2021). This disproportionate burden, which also affects men but to a lesser extent, is evident in various contexts, manifesting as sexual violence in high school (14% of girls vs. 3% of boys) (Hébert et al., 2019) and sexual harassment in the workplace (16% of women vs. 8% of men) (Burczycka, 2021). This raises a critical question: in a context of a strong welfare state with high female labour force participation and education attainment, how does this persistently high prevalence of violence shape women’s economic outcomes?

This study investigates the relationship between sexual and physical violence over the life course and LFP among both women and men in Canada, using nationally representative survey data linked to tax data. Given the high educational attainment and labour force participation of Canadian women (Moyser, 2019; Pelletier et al., 2019), we hypothesize that women who report no lifetime violence exposure will have LFP rates and income approaching those of unexposed men. Second, we hypothesize that violence exposure creates a distinct disadvantage: exposed women will be (1) less likely to be economically active and (2) more likely to be employed in lower-paying jobs or sectors, compared to their unexposed counterparts.

## Methods

### Data Source and Study Sample

This study analyzed data from the 2018 Canadian Survey on Safety in Public and Private Spaces (SSPPS) (Statistics Canada, 2018), a cross-sectional survey conducted by Statistics Canada between April and December 2018 and linked to respondents’ tax records for the 2017 tax year, with a filing due date of April 2018 (Canada Revenue Agency, 2021). The 2018 cycle is the most currently available (as of the time of writing), and is, to our knowledge, the only Statistics Canada survey to comprehensively assess both exposure to lifetime violence and LFP. The SSPPS collected data from a stratified, multistage probability sample of individuals aged 15 and older residing in Canada’s ten provinces, excluding those living in its three Territories, in institutions or on reserves. Data were gathered in English and French via telephone and online questionnaires. The full SSPPS 2018 sample consists of 45,893 respondents (Statistics Canada, 2024). The survey achieved a response rate of 43.1%, comparable to other Statistics Canada surveys from similar periods such as the General Social Survey (Government of Canada, 2019) and the Canadian Community Health Survey (Anderson et al., 2025).

This analysis focuses on a subset of the SSPPS respondents. We first restricted the sample to respondents of working age (18 to 64). From this group, we then included only those who provided valid responses to the LFP and violence questions described below. Based on these inclusion criteria, our final analytic sample consists of approximately 30% of the full SSPPS sample.

### Exposure

The primary exposure variable was lifetime exposure to violence, spanning childhood and adulthood. Survey items used to measure violence are summarized in Supplementary Table 1. The SSPPS defines childhood violence as self-reported sexual or severe physical victimization by an adult before age 15. This definition does not include measures of neglect or common corporal punishment (e.g., spanking). Adulthood violence included self-reported physical or sexual violence after the age of 15 years, excluding acts perpetrated by an intimate partner. IPV in adulthood was measured using separate items describing coercive, physical, and sexual behaviours committed by current or former intimate partners. Our main exposure variable was defined as violence occurring in either childhood or adulthood, excluding exposures reported in the 12 months preceding the survey, to ensure that exposure (violence) preceded the outcomes (past-year income and labour force participation).

### Outcomes

LFP was determined using a sequence of SSPPS survey questions. All respondents were first asked: (1) “Last week, did you work at a job or business?” Those who answered “No” were then asked: (2) “Last week, did you have a job or business from which you were absent?” If the response was still negative they were asked a final question: (3) “In the past 12 months, did you work at a job or business?” Respondents who answered “Yes” to any of these three questions were classified as economically active in the past year. Conditioning on those who worked in the past year, we examined part- and full-time status. Full-time versus part-time employment was derived from responses to the item: “On average, how many (paid) hours did you usually work per week (excluding overtime)?” Consistent with Statistics Canada definitions (Statistics Canada, 2011), respondents who reported 30 or more hours per week were categorized as full-time; those reporting fewer than 30 hours were categorized as part-time. Those who answered “No” to all three items above were classified as economically inactive. Annual personal income was derived from administrative tax filing records linked to the SSPPS and reflects total reported income from all sources in 2018, the calendar year preceding the survey.

### Analyses

We first described the demographic, socioeconomic, and health characteristics of the analytic sample, stratified by past-year LFP. We then examined self-reported experiences of violence by gender across childhood, adulthood, and intimate partnerships, and assessed how patterns of exposure varied by economic activity. To explore associations between violence exposure and labour market outcomes, we compared employment status across gender and violence exposure. Among respondents not engaged in paid work in the past year, we analyzed reasons for inactivity, including health issues, caregiving responsibilities, retirement, and job search or education.

Among those employed in the past year, we examined part-time versus full-time status, as well as the distribution of workers across industry sectors and occupational categories by gender and violence exposure. Industry sectors were classified using the 2017 North American Industry Classification System (NAICS) (Statistics Canada, 2016). To comply with disclosure vetting rules while maintaining interpretability, NAICS codes were collapsed into broader categories based on conceptual similarity (e.g., grouping natural resources, agriculture, and extractive sectors; or combining education, law, and social services into a single occupational category). We also explored how violence exposure relates to skill level by examining the distribution of workers across employment sectors, stratified by educational attainment (secondary or less vs. more than secondary). Chi-squared tests were used to assess group differences in sectoral distribution across three comparisons: (1) exposed vs. unexposed individuals within each gender, (2) men vs. women overall, and (3) exposed vs. unexposed individuals regardless of gender.

Finally, we used inverse probability weighted regression adjustment (IPWRA) to estimate the average treatment effect (ATE) of violence exposure on the log-odds of past-year economic inactivity, and on specific reported reasons for inactivity. ATE measures the average difference in outcomes between a group that receives a treatment (or intervention) and a group that does not after adjusting for the influence of identified confounders (Angrist & Pischke, 2009). All models were adjusted for a set of demographic and socioeconomic covariates and used unexposed men as the reference group. We use unexposed men as the comparison group because they provide a counterfactual representing individuals not subject to violence-related constraints or barriers to LFP, enabling clearer estimation of the incremental impact of violence on women’s economic inactivity after adjusting for confounders. Covariate selection was informed by the literature and included age, place of birth (i.e., born in Canada vs. not), visible minority status (Government of Canada, 2021b), marital status, presence of minors in the household, educational attainment, and self-rated general health.

In observational studies like ours, where randomized controlled trials (RCTs) are not feasible, methods such as IPWRA are used to achieve covariate balance. Covariate balance ensures that differences in outcomes can be attributed to the exposure of interest rather than confounding variables. IPWRA combines matching and regression adjustment by applying weights to each observation, aligning the distribution of covariates across groups without discarding data (Chesnaye et al., 2022; Wooldridge, 2007). This dual approach not only improves balance but also enhances robustness to model misspecification, compared to methods such as propensity score matching, enabling accurate estimation of ATEs (Imai & Ratkovic, 2014; Wooldridge, 2007) (Wooldridge, 2007; Imai and Ratkovic, 2014). By using IPWRA, this study examines the relationship between gender, violence exposure, and past-year economic inactivity, accounting for potential confounding factors while allowing for more precise group comparisons.

To identify nuanced income patterns that aggregated models might obscure, we descriptively examined mean personal income across employment sectors and education levels. This exploratory analysis is included to highlight potential wage disparities and generate hypotheses for future research. Formal statistical testing was not conducted due to the large number of subgroup combinations (80 cells across 10 sectors, 4 gender/exposure groups, and 2 education levels), which would inflate the risk of Type I errors.

All descriptive and multivariable analyses incorporated survey and bootstrap weights provided by Statistics Canada to address nonresponse and ensure representativeness of the Canadian population. Statistical significance was assessed at the 5% level. Analyses were conducted using Stata 17 (StataCorp, 2021).

### Sensitivity Analyses

To assess the robustness of our findings and test the stability of estimates under alternative definitions of violence exposure, we conducted a series of sensitivity analyses using three additional exposure indicators beyond our main specification. While our primary models examined the effect of lifetime violence exposure excluding incidents in the past 12 months, we also estimated separate IPWRA models for: (1) any lifetime violence exposure, regardless of timing, (2) exposure to violence occurring in both childhood and adulthood, and (3) recent exposure within the past 12 months.

## Results

### Sample Characteristics

Nearly two-thirds of women (64.5%) and three in five men (59.1%) reported experiencing at least one form of physical or sexual violence over their lifetime. Prevalence was also slightly higher among those who had worked in the past year (62.0%) compared to those who had not (60.8%). Differences in violence exposure by gender and past-year LFP were both statistically significant (p-value, p<0.001). Detailed breakdowns by violence type and subgroup are presented in Supplementary Tables 2 and 3.

A total of 85.9% of respondents reported economic activity in the past year. The median age was 42 years (IQR: 32–54), and the sample was nearly evenly split by gender (49.4% women and 50.6% men). The majority of respondents were born in Canada (74.5%), with 25.0% born outside Canada and 0.5% not stating their place of birth. Visible minorities comprised 21.3% of the sample, while 77.7% identified as non-visible minorities. These distributions are broadly consistent with national population patterns, where approximately one in four Canadians is foreign-born and just over one in five identifies as a visible minority (Statistics Canada, 2022, 2023), and the sample’s age and gender composition is comparable to census estimates, though slightly older than the national median age of 41 (Statistics Canada, 2017).

The majority (75.9%) of respondents were married or in common-law relationships, and 19.0% were single, separated, divorced, or widowed. Approximately 38.7% of respondents reported having children under 17 in their household. Educational attainment was high, with 34.9% holding a bachelor’s degree or higher, 36.4% having a diploma or trade certificate, and 6.3% not completing high school. Self-rated health was overwhelmingly positive, with 90.9% of respondents rating their health as good to excellent.

When aggregated across all types of lifetime violence exposure, there were notable differences in employment status between men and women (Table 2). Among unexposed men, nearly all (90.1%) were economically active. For women, economic activity was slightly higher among those exposed to violence (82.0%) than unexposed women (80.3%). However, while differences between men and women by exposure status were not statistically significant, the gap in economic activity between men and women was significant (90.1% vs. 81.4%; p < 0.001).

**Table 1:** Descriptive Characteristics of the Study Cohort.

| **Table 1: Descriptive Characteristics of the Study Cohort** |  |
| --- | --- |
|  | Proportion |
| **Lifetime Exposure by Gender (Women)** |  |
| Exposed | 64.5% |
| Unexposed | 35.5% |
| **Lifetime Exposure by Gender (Men)** |  |
| Exposed | 59.1% |
| Unexposed | 40.9% |
| **Past-year Economic Activity** |  |
| Worked at a job or business in the past year | 85.9% |
| Did not work at a job or business in the past year | 14.1% |
| **Age** |  |
| Median [IQR] | 42 [32-54] |
| **Gender** (Column [Col] %) |  |
| Woman | 49.4% |
| Man | 50.6% |
| **Born In Canada** (Col %) |  |
| Yes | 74.5% |
| No | 25.0% |
| Not Stated | 0.5% |
| **Visible Minority** (Col %) |  |
| No | 77.7% |
| Yes | 21.3% |
| Not Stated | 1.0% |
| **Marital Status (Col %)** |  |
| Married | 58.2% |
| Common-law | 17.7% |
| Widowed | 0.2% |
| Separated | 2.0% |
| Divorced | 2.9% |
| Single (Never married) or Not Stated | 19.0% |
| **Respondents Children Who Are Minors (Age < 17) Present in Household (Col %)** |  |
| None or Not Stated | 61.3% |
| Yes | 38.7% |
| **Highest Educational Qualification Attained (Col %)** |  |
| Less than Highschool or Not Stated | 6.3% |
| High School | 22.3% |
| Diploma or Trade Certificate | 36.4% |
| Bachelors and Higher | 34.9% |
| **Self-rated General Health (Col %)** |  |
| Fair, Poor, or Not Stated | 9.1% |
| Good to Excellent | 90.9% |

**Table 2:** Employment Status by Lifetime Violence Exposure and Gender.

| Lifetime Violence Exposure and Gender [Row %] | Did Not Work<br>Past Year | Worked Past<br>Year | Total |
| --- | --- | --- | --- |
| Unexposed Men | 10.0% | 90.1% | 100.0% |
| Exposed Men | 9.4% | 90.7% | 100.0% |
| Unexposed Women | 19.7% | 80.3% | 100.0% |
| Exposed Women | 18.0% | 82.0% | 100.0% |
| Total | 14.1% | 85.9% | 100.0% |
| Chi-square p-value < 0.01 (all associations) |  |  |  |

We used IPWRA models to estimate the ATE of lifetime violence exposure,excluding incidents in the past 12 months, on past-year economic inactivity, using unexposed men as the reference group (Table 3). Among men, the ATE was small and not statistically significant (0.021; 95% CI: –0.009, 0.050). For women, both exposed and unexposed groups showed significantly higher probabilities of inactivity compared to unexposed men (ATE: 0.096; 95%CI: [0.075, 0.117] for unexposed women and 0.092; 95%CI: [0.066, 0.118] for exposed women), but the difference between exposed and unexposed women was not statistically significant.

**Table 3:**
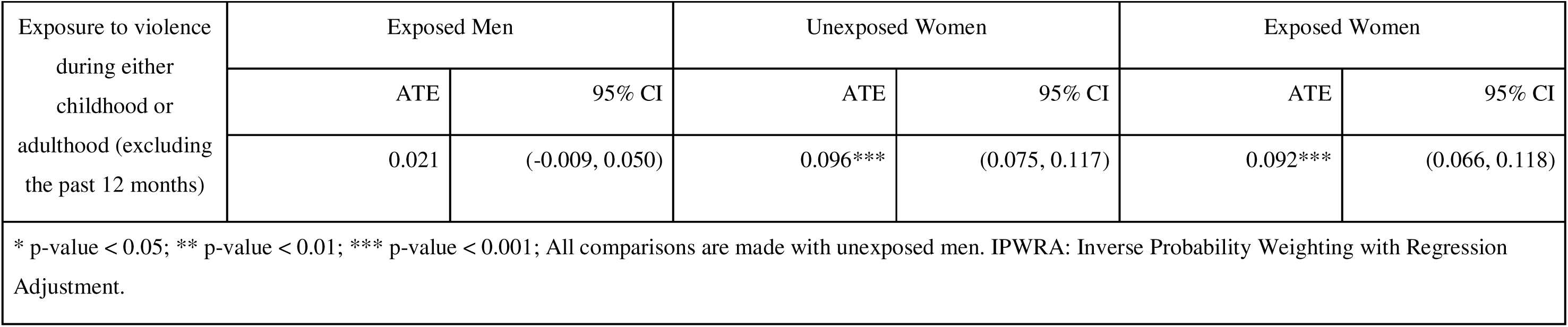
IPWRA Results for Log Odds of Past-year Economic Inactivity Stratified by Type of Exposure.

To examine potential mechanisms, we analyzed reasons for employment interruption among those who did not work in the past year. Descriptive analysis (Table 4) revealed strong differences for men compared to women (p<0.001): women were much more likely to cite family and household responsibilities, while men more often reported retirement. Health-related inactivity was more common among exposed men and women relative to their unexposed counterparts.

**Table 4:** Past-Year Economic Inactivity by Reason and Exposure to Violence and Gender.

|  |  |  |  |  |  |
| --- | --- | --- | --- | --- | --- |
|  | **Unexposed**<br>**Men** | **Exposed**<br>**Men** | **Unexposed**<br>**Women** | **Exposed**<br>**Women** | **Total** |
| Job Search | 9.4% | 7.0% | 3.1% | 2.4% | 4.5% |
| Education | 8.2% | 10.9% | 9.9% | 8.9% | 9.5% |
| Family and Household Responsibilities | 6.1% | 7.0% | 43.9% | 43.5% | 30.9% |
| Retirement | 53.5% | 43.8% | 32.6% | 26.9% | 35.6% |
| Health Issues | 16.8% | 25.3% | 7.1% | 12.6% | 14.4% |
| Other Or Missing | 5.7% | 5.5% | 3.2% | 5.6% | 5.0% |
| Total | 100.0% | 100.0% | 100.0% | 100.0% | 100.0% |

Adjusted IPWRA models (Table 5) showed that violence exposure among women was modestly but significantly associated with higher probabilities of retirement (ATE: 0.015; 95% CI: 0.000, 0.031) and health-related inactivity (ATE: 0.009; 95% CI: 0.000, 0.017), relative to unexposed men. No significant associations were found for job search or education-related inactivity. Among men, ATEs remained small and non-significant across all reasons for inactivity. While some effects among women reached statistical significance, they were small in magnitude (under two percentage points), and confidence intervals overlapped between exposed and unexposed individuals within gender, indicating no clear within-gender differences by exposure status.

**Table 5:**
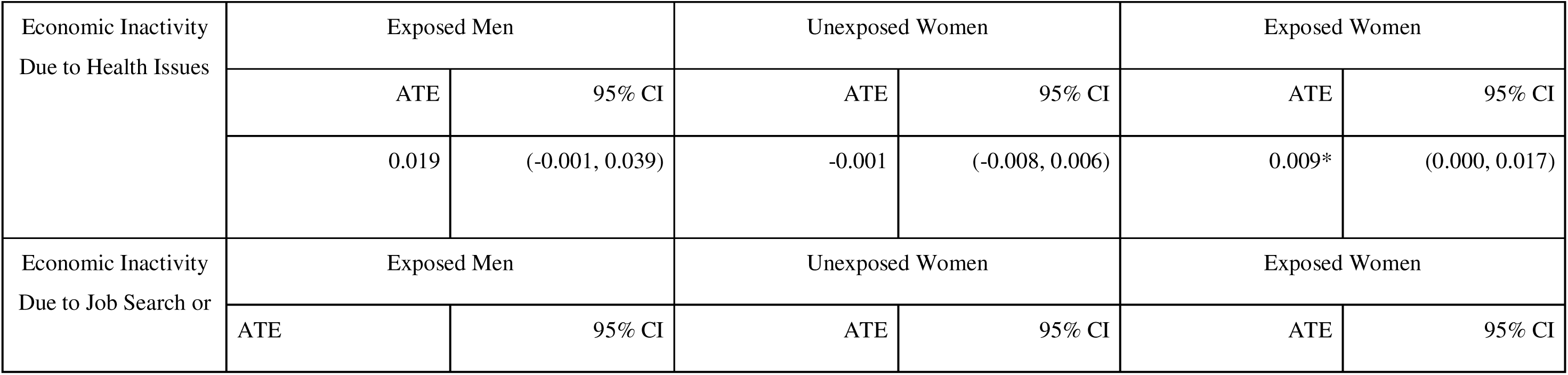
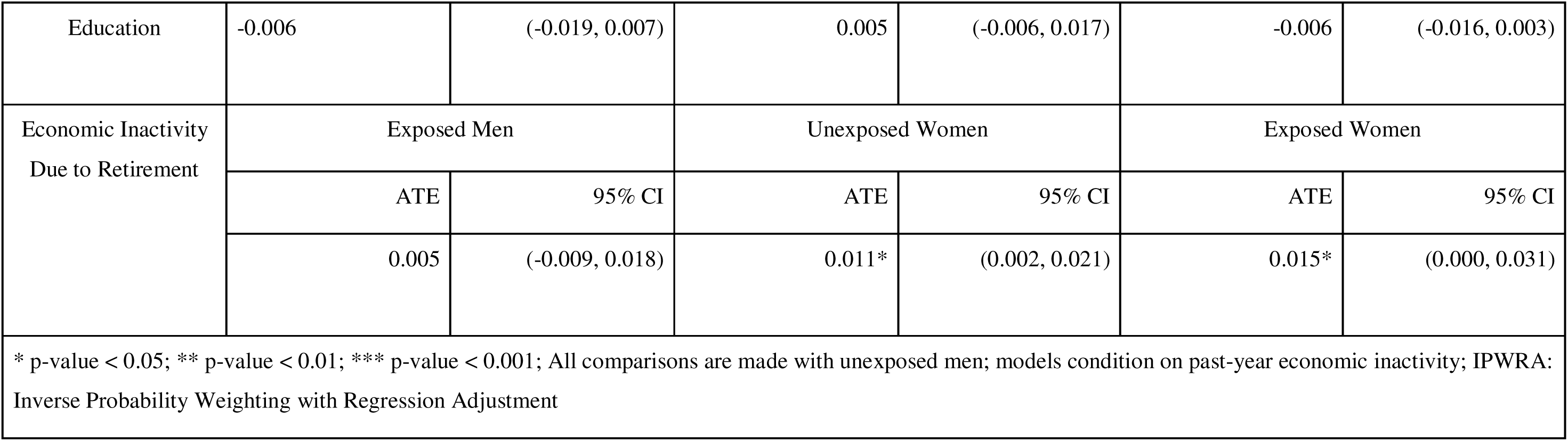
IPWRA Estimates of Log-Odds for Past-Year Economic Inactivity by Reason, Exposure to Violence, and Gender.

|  |  |  |  |  |  |  |
| --- | --- | --- | --- | --- | --- | --- |
| Economic Inactivity<br>Due to Health Issues | Exposed Men |  | Unexposed Women |  | Exposed Women |  |
|  | ATE | 95% CI | ATE | 95% CI | ATE | 95% CI |
|  | 0.019 | (-0.001, 0.039) | -0.001 | (-0.008, 0.006) | 0.009* | (0.000, 0.017) |
| Economic Inactivity<br>Due to Job Search or | Exposed Men |  | Unexposed Women |  | Exposed Women |  |
|  | ATE | 95% CI | ATE | 95% CI | ATE | 95% CI |
| Education | -0.006 | (-0.019, 0.007) | 0.005 | (-0.006, 0.017) | -0.006 | (-0.016, 0.003) |
| Economic Inactivity<br>Due to Retirement | Exposed Men |  | Unexposed Women |  | Exposed Women |  |
|  | ATE | 95% CI | ATE | 95% CI | ATE | 95% CI |
|  | 0.005 | (-0.009, 0.018) | 0.011* | (0.002, 0.021) | 0.015* | (0.000, 0.031) |
| * p-value < 0.05; ** p-value < 0.01; *** p-value < 0.001; All comparisons are made with unexposed men; models condition on past-year economic inactivity; IPWRA: Inverse Probability Weighting with Regression Adjustment |  |  |  |  |  |  |

Part-time versus full-time status among those employed in the past year showed minimal differences by violence exposure among women. In Table 6, part-time employment rates were virtually identical for exposed and unexposed women (28.5% vs. 28.7%), suggesting that lifetime violence exposures did not significantly influence whether women worked part-time. Among men, however, part-time employment was slightly more common among those exposed to violence (12.0% vs. 11.1%). While these differences were statistically significant, their small magnitude limits their substantive interpretation.

**Table 6:** Part-time/Full-time status by Lifetime Violence Exposure and Gender.

|  |  |  |  |  |  |
| --- | --- | --- | --- | --- | --- |
|  | Unexposed Men | Exposed Men | Unexposed Women | Exposed Women | Total |
| Part-Time | 11.1% | 12.0% | 28.7% | 28.5% | 19.5% |
| Full Time | 87.7% | 86.8% | 68.9% | 70.3% | 79.0% |
| Not Stated | 1.2% | 1.1% | 2.5% | 1.2% | 1.4% |
| Total | 100.0% | 100.0% | 100.0% | 100.0% | 100.0% |
| Chi-square p-value < 0.01; Chi-square p-value comparing (1) unexposed men to exposed men, and (2) unexposed women to unexposed women was also <0.01 |  |  |  |  |  |

Differences in industry sector (NAICS) appeared to be primarily driven by gender rather than violence exposure (Table 7). Regardless of exposure, men were more concentrated in manufacturing, construction, and extractive industries, while women were more likely to work in health, education, and service sectors. Comparisons within gender showed only modest differences by exposure status. For example, exposed men were slightly less likely than unexposed men to work in manufacturing and construction (21.5% vs. 25.3%) and slightly more likely to be employed in professional services (15.7% vs. 14.7%) and public administration (8.1% vs. 5.7%). Among women, those exposed to violence were more likely to work in educational services (13.2% vs. 10.7%) and less likely to work in health services (21.7% vs 24.9%) compared with those unexposed to violence.

**Table 7:** Labour Force Participation by Sector, Lifetime Violence Exposure, and Gender (Among those who worked in the last year)

|  | **Unexposed**<br>**Men** | **Exposed**<br>**Men** | **Unexposed**<br>**Women** | **Exposed**<br>**Women** | **Total** |
| --- | --- | --- | --- | --- | --- |
| Agriculture, Fishing/Hunting, Forestry,<br>and Extractive | 6.1% | 4.8% | 1.8% | 1.8% | 3.7% |
| Manufacturing & Construction | 25.3% | 21.5% | 7.8% | 6.4% | 15.5% |
| Wholesale and Retail Trade | 11.4% | 13.0% | 14.8% | 13.0% | 12.9% |
| Transportation and Warehousing | 7.7% | 6.6% | 2.8% | 2.5% | 5.0% |
| Information, Cultural, and Financial<br>Services | 8.5% | 7.3% | 6.9% | 7.8% | 7.7% |
| Professional and Business Services | 14.7% | 15.7% | 12.0% | 12.7% | 14.0% |
| Educational Services | 5.9% | 6.8% | 10.7% | 13.2% | 9.2% |
| Health Services | 4.4% | 4.5% | 24.9% | 21.7% | 13.1% |
| Public Administration | 5.7% | 8.1% | 6.5% | 8.2% | 7.4% |
| Other Services | 10.2% | 11.7% | 11.8% | 12.6% | 11.7% |
| **Total** | 100.0% | 100.0% | 100.0% | 100.0% | 100.0% |
| NAICS: North American Industrial Classification System (2017) (10 categories); Chi-square p-value < 0.01 |  |  |  |  |  |

Chi-squared tests indicated statistically significant differences in sectoral (NAICS) distributions across all comparisons, by violence exposure within gender, by gender regardless of exposure, and by exposure regardless of gender (all p < 0.01). However, given the number of categories, these tests are highly sensitive to small differences and may not always indicate substantively meaningful variation.

When considering skill levels across industries (Table 8), differences by violence exposure were small and inconsistent. In the information, cultural, and financial services sector, 85.5% of exposed men had more than a secondary education, compared to 86.8% of unexposed men.

**Table 8:** Distribution of Skill by Sector, Lifetime Violence Exposure, and Gender among those who worked in the past year.

|  | **Unexposed**<br>**Men** | **Exposed**<br>**Men** | **Unexposed**<br>**Women** | **Exposed**<br>**Women** | **Total** |
| --- | --- | --- | --- | --- | --- |

|  | *Secondary or less*<br>*(Row total)* | *More than secondary*<br>*(Row total)* | *Column Total* | *Secondary or less*<br>*(Row total)* | *More than secondary*<br>*(Row total)* | *Column Total* | *Secondary or less*<br>*(Row total)* | *More than secondary*<br>*(Row total)* | *Column Total* | *Secondary or less*<br>*(Row total)* | *More than secondary*<br>*(Row total)* | *Column Total* | *Secondary or less*<br>*(Row total)* | *More than secondary*<br>*(Row total)* | *Column Total* |
| --- | --- | --- | --- | --- | --- | --- | --- | --- | --- | --- | --- | --- | --- | --- | --- |
| Agriculture, Fishing/Hunting, Forestry, and Extractive | 52.6% | 47.7% | 6.1% | 39.5% | 60.5% | 4.8% | 44.9% | 55.1% | 1.8% | 33.5% | 66.5% | 1.8% | 43.7% | 56.3% | 3.7% |
| Manufacturing & Construction | 38.7% | 61.4% | 25.3% | 34.1% | 65.9% | 21.5% | 35.3% | 65.0% | 7.8% | 32.5% | 67.5% | 6.4% | 35.6% | 64.4% | 15.5% |
| Wholesale and Retail Trade | 39.5% | 60.5% | 11.4% | 41.2% | 58.9% | 13.0% | 41.7% | 58.4% | 14.8% | 39.6% | 60.4% | 13.0% | 40.5% | 59.6% | 12.9% |
| Transportation and Warehousing | 49.2% | 50.8% | 7.7% | 45.4% | 54.6% | 6.6% | 43.4% | 57.4% | 2.8% | 35.4% | 65.0% | 2.5% | 44.9% | 55.2% | 5.0% |
| Information, Cultural, and Financial Services | 13.4% | 86.8% | 8.5% | 14.6% | 85.5% | 7.3% | 23.8% | 76.5% | 6.9% | 16.5% | 83.5% | 7.8% | 16.2% | 83.8% | 7.7% |
| Professional and Business Services | 18.2% | 81.8% | 14.7% | 17.4% | 82.7% | 15.7% | 16.1% | 83.9% | 12.0% | 20.3% | 79.8% | 12.7% | 18.2% | 81.9% | 14.0% |
| Educational Services | 10.8% | 89.2% | 5.9% | 8.9% | 91.1% | 6.8% | 11.3% | 88.7% | 10.7% | 10.7% | 89.4% | 13.2% | 10.3% | 89.7% | 9.2% |
| Health Services | 10.3% | 90.0% | 4.4% | 13.7% | 86.3% | 4.5% | 15.9% | 84.1% | 24.9% | 11.9% | 88.1% | 21.7% | 13.2% | 86.8% | 13.1% |
| Public | 16.7% | 83.3% | 5.7% | 14.1% | 85.9% | 8.1% | 13.9% | 86.1% | 6.5% | 11.6% | 88.4% | 8.2% | 13.6% | 86.4% | 7.4% |
| Administration |  |  |  |  |  |  |  |  |  |  |  |  |  |  |  |
| Other Services | 35.4% | 64.6% | 10.2% | 34.6% | 65.5% | 11.7% | 31.0% | 69.2% | 11.8% | 40.7% | 59.4% | 12.6% | 36.1% | 63.9% | 11.7% |
| **Total** | 30.7% | 69.3% | 100.0% | 27.7% | 72.3% | 100.0% | 24.2% | 75.8% | 100.0% | 22.6% | 77.4% | 100.0% | 26.2% | 73.8% | 100.0% |
| NAICS: North American Industrial Classification System (2017) (10 categories) |  |  |  |  |  |  |  |  |  |  |  |  |  |  |  |

We descriptively examined mean personal income across employment sectors by education level and lifetime exposure to violence (Table 9). Among women, income differences by exposure status were inconsistent in direction and varied by skill level. Overall, exposed women earned slightly more than unexposed women ($53,787 vs. $52,441), driven primarily by those with more than a secondary education ($59,393 vs. $56,439). In contrast, among women with only secondary education or less, the trend was reversed and more pronounced: exposed women earned substantially less than their unexposed counterparts ($34,604 vs. $39,913).

**Table 9:**
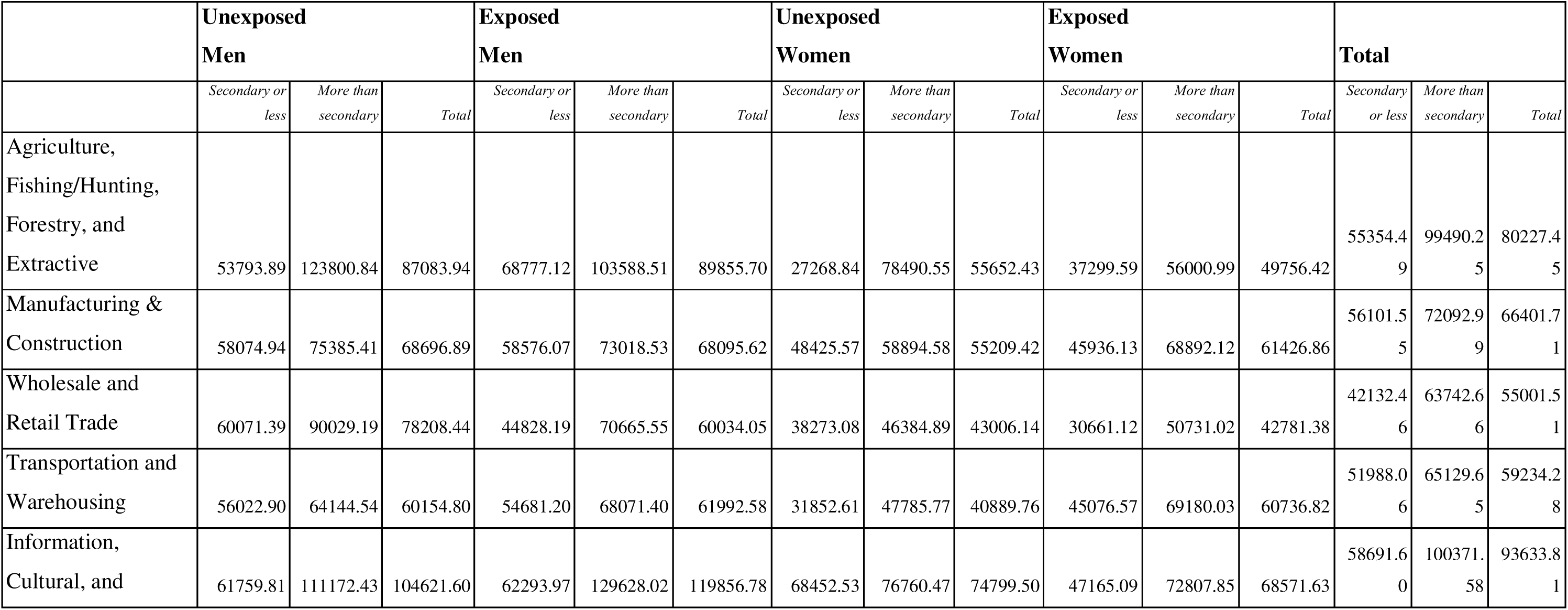
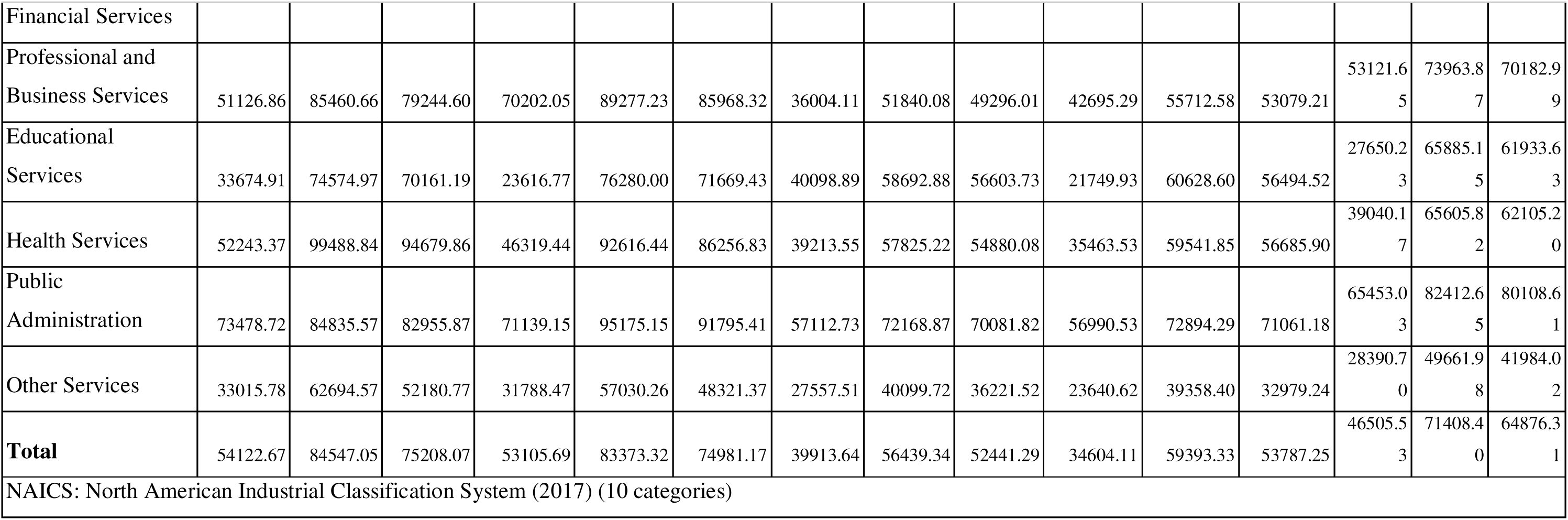
Mean wage by Sector, Skill, Lifetime Violence Exposure, and Gender.

These patterns were more consistent among men. Across all education levels, exposure to violence was associated with lower average income. Overall, exposed men earned $74,981 compared to $75,208 for unexposed men. Among men with secondary education or less, exposed men earned $53,106 vs. $54,123; among those with higher education, exposed men earned $83,373 vs. $84,547.

### Supplementary Analyses

We repeated the IPWRA models using alternative definitions of violence exposure, as detailed in the Methods section, and found results consistent with the main analysis (Supplementary Table 4). Among men, ATEs remained small and statistically non-significant across all definitions. For women (compared to unexposed men), estimates were similar in magnitude regardless of the exposure window, and differences between exposed and unexposed women were not statistically significant. The highest ATE was observed among women exposed in the past 12 months (ATE: 0.132; 95% CI: 0.056, 0.207).

## Discussion

Using nationally representative data, we examined whether lifetime exposure to violence was associated with past-year economic inactivity, part-time work, occupational and sectoral sorting, and income differences. This study fills a critical gap by exploring the labour market impacts of violence in Canada, a high-income country with comparatively strong social protections. To our knowledge, this is the first study to investigate the relationship between violence, labour force participation, and income using the inaugural SSPPS wave, offering novel insights into how violence may influence economic outcomes in a high-income, welfare-state context. Our approach, which combines survey and administrative tax data and disaggregates employment outcomes beyond simple labour force participation (e.g., by occupation and skill level), offers a methodological template that can be applied in other settings, including LMICs undergoing rapid economic and social change. We found limited evidence that violence exposure reduces women’s labour force participation in this setting.

Economic inactivity rates were similar between exposed and unexposed women, though exposed women were slightly more likely to report health-related inactivity and early retirement. These differences were small in magnitude but statistically significant. The more pronounced distinctions emerged in how survivors participated in the labour market: exposed women were more likely to work part-time and to be concentrated in care-oriented sectors such as education and community services, though not necessarily in lower-skilled roles.

Income differences by exposure status varied by sector and education level, with no consistent pattern. In some sectors, exposed women reported higher incomes compared to their unexposed counterparts; in others, less. These findings suggest that violence may shape women’s economic outcomes less by reducing overall participation or income, and more through occupational sorting, part-time work, and sectoral clustering, though these interpretations remain tentative, as they are based on descriptive analyses and warrant further investigation. While men’s patterns were generally weaker and more inconsistent, they help underscore the gendered nature of these associations.

Together, these results suggest that in the Canadian context, the economic impacts of violence are complex and, critically, do not include the large-scale labour force exits seen in other settings. Our finding that lifetime violence exposure had no significant impact on women’s overall LFP suggests that Canada’s social safety nets (e.g., universal healthcare, income supports) may act as a crucial buffer that mitigates the most catastrophic economic consequences of violence and prevents survivors from being forced out of the workforce entirely.

Our finding that violence exposure was not associated with a significant reduction in women’s labour force participation contrasts with evidence from other contexts. Henkhaus (2022), using US data, found that childhood sexual violence was associated with a 4.6 percentage point drop in full-time employment among women. Ouedraogo and Stenzel (2021) similarly reported that a one percentage-point increase in IPV prevalence corresponded to an 8% decline in district-level economic activity across Sub-Saharan Africa, largely driven by reduced LFP among women. By contrast, we observed no significant difference in labour force participation between exposed and unexposed women in Canada (82.0% vs. 80.3%), with adjusted models suggesting that gender, not violence exposure, was the primary driver of economic inactivity. Our results align more closely with those of Gedikli et al. (2023), who found a 6.2 percentage point increase in LFP among Turkish women exposed to IPV, potentially reflecting survival-driven labour market entry or coercive economic control. These differences highlight the importance of national context—including labour market structures, income levels, and especially the strength of social protections, in shaping how violence influences employment. Notably, we did observe a clearer income gap among women with secondary education or less: unexposed women reported roughly $40,000 on average, compared to $35,000 among those exposed. Though exploratory, this pattern suggests that education may amplify the economic vulnerability associated with violence exposure and warrants deeper investigation in future work.

Our descriptive findings also suggest possible occupational sorting among women based on lifetime exposure to violence. Exposed women appeared slightly more concentrated in sectors such as education and cultural services, while being comparatively underrepresented in health services. However, these patterns are preliminary and should be interpreted with caution. Without adjusted models, it is difficult to disentangle the influence of violence exposure from broader gendered labour market dynamics, particularly given the well-documented tendency for women, regardless of violence history, to be overrepresented in these sectors (Bamieh & Ziegler, 2024; Morales & Marcén, 2024). At this stage, the observed differences are better understood as gender effects rather than definitive consequences of violence. Still, the possibility that violence exposure may shape occupational decisions or constraints remains underexplored. Given the limited empirical literature in this area, further research is warranted to examine how violence contributes to occupational sorting independently of gender, ideally using longitudinal or causal designs. These findings also suggest that researchers should further investigate how the welfare state may act as a buffer to moderate earnings differentials due to violence.

Although overall rates of labour force participation did not differ significantly by violence exposure, our adjusted models revealed that exposed women were slightly more likely to cite health problems and early retirement as reasons for economic inactivity. While small in magnitude, these differences align with well-established pathways linking violence to chronic physical and mental health conditions. As noted earlier, survivors of violence are at increased risk for chronic pain (Wuest et al., 2008), disability (Ballan et al., 2022), PTSD (Dutton et al., 2006; Iverson et al., 2017), depression (Ahmadabadi et al., 2020; Devries et al., 2013; Spencer et al., 2023), and anxiety (Ahmadabadi et al., 2020)–all of which can reduce one’s capacity to remain employed, productive at work or delay retirement. The modest increase in health-related and early retirement inactivity among exposed women in our study offers empirical support for this theorized mechanism. While our cross-sectional data cannot confirm causality, these patterns suggest that violence exposure may shape not only whether women work, but under what conditions they exit the labour market.

To better understand these dynamics, future research should leverage longitudinal data to assess the long-term economic consequences of violence. Tracking income trajectories is essential to distinguish between temporary setbacks and permanent labour market exits, and to establish causal directionality. Crucially, such analyses could also disentangle the mechanisms of exit, determining whether health-related inactivity reflects solely physical incapacity or a pathway for survivors not yet eligible for retirement to access disability supports. Monitoring these associations over time will clarify whether social protections continue to buffer survivors against economic scarring amidst broader instability, or if disadvantages persist.

This study has several notable strengths. It draws on a nationally representative sample from the 2018 SSPPS, enabling generalizability to the broader Canadian population and to similar high-income contexts with robust social support systems. The dataset includes detailed measures of lifetime exposure to violence, allowing for analyses across multiple forms of abuse and indicators for LFP and income, including violence in both childhood and adulthood. This study also analyzes the impact of violence among both men and women. The use of IPWRA strengthens internal validity by addressing confounding from observed variables. The study also benefits from linkage to administrative tax data, providing more reliable income estimates than self-reported earnings, and from a comprehensive approach that stratifies economic outcomes between men and women, and by sector, occupation, and skill level.

However, several limitations should be noted. First, the cross-sectional nature of the data precludes causal inference. While associations are reported, we cannot determine whether violence exposure caused the observed economic outcomes and sectoral sorting or whether pre-existing vulnerabilities increased the risk of violence. This limitation is partially mitigated by our decision to exclude past-year exposures from our definition of violence, ensuring that the exposures temporally precede the measured outcomes. Second, although IPWRA adjusts for a rich set of observed confounders, unmeasured variables, such as, access to social services, may still bias the estimated relationships. Third, violence exposure is self-reported and may be underreported due to stigma, fear, or recall bias. This likely results in conservative estimates of both the prevalence and effects of violence, although the detailed, behaviorally specific nature of the SSPPS questions may improve disclosure relative to other surveys. Fourth, while the use of linked tax data improves income measurement, it does not capture informal or unreported income. More importantly, the data linkage provided only aggregated total income, which includes non-wage sources like social assistance and pensions. This may obscure the true impact of violence on earnings. This could also bias the comparison if survivors are systematically more likely to receive such transfers, as this “cushioning” would make their total income appear artificially similar to the unexposed group. Moreover, both LFP and income were measured only for the past year, providing a limited snapshot of survivors’ economic conditions. As discussed earlier, future research could link SSPPS respondents to a broader window of tax records to assess longer-term impacts, such as persistent income loss or delayed workforce entry. Finally, the survey’s 43% response rate, though addressed through weighting and still higher than waves of most other Statistics Canada surveys (Government of Canada, 2019, 2021a), raises the possibility of nonresponse bias. The SSPPS weights help align the sample with national census distributions on key sociodemographic characteristics such as age, gender, immigrant status, and visible minority status, which improves representativeness. However, these adjustments cannot fully account for the potential underrepresentation of individuals who are most marginalized or most affected by violence, for example, 80% of male and 86% of female prisoners have reported experiencing childhood violence (Sheahan, 2023), but this population is not captured in the SSPSS (Government of Canada, 2024). As a result, our findings may underestimate the true prevalence and economic consequences of violence in Canada.

These findings highlight the nuanced ways in which violence exposure influences labour market experiences in Canada. While overall economic inactivity did not differ substantially by exposure status, women survivors did face increased risks of health-related exits and early retirement, pointing to the far-reaching toll of violence on well-being and labour force participation. Although we did not find that violence was associated with large-scale labour force exit as in other contexts, this does not mean that violence is without economic cost; rather, it suggests the impacts are more complex and raises a crucial question for future research: to what extent, and through which mechanisms, do social and labour safety nets (e.g., universal healthcare, income supports) act as a buffer against these more catastrophic economic consequences. Furthermore, our findings for men were less clear and likely underestimated. Given that surveys like the SSPPS exclude highly impacted, marginalized groups (such as incarcerated men, among whom violence exposure is exceptionally high), a mixed-methods approach is essential. Qualitative work, in particular, is needed to understand the specific economic trajectories and support needs of male survivors, whose experiences remain obscured in this quantitative data.

## Data Availability

The datasets analysed during the current study are not publicly available. They are housed securely at the McMaster University Statistics Canada Research Data Centre (RDC) and are protected by federal law (the Statistics Act). Access is strictly limited to authorized researchers named on the project contract who have undergone the requisite security vetting.

## Acknowledgements

We acknowledge the support of the Li Ka Shing Knowledge Institute of Unity Health Toronto. We acknowledge the support of the Canadian Research Data Centre Network without whom the data underlying this work would not be possible. This research was also partially supported by the Fondation Botnar and the Institute of Gender and the Economy.

## Supplemental Materials

**Supplementary Table 1:**
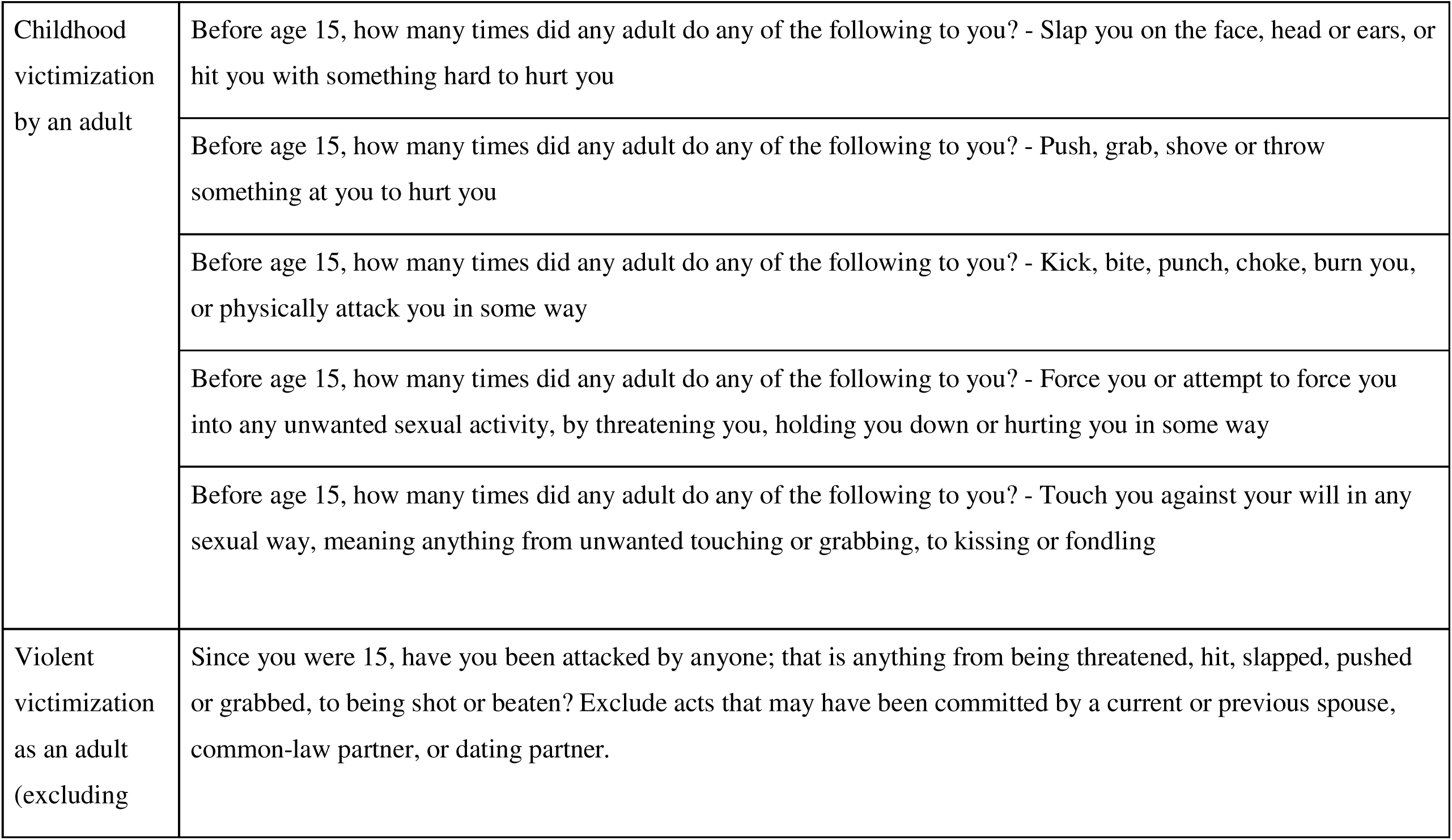
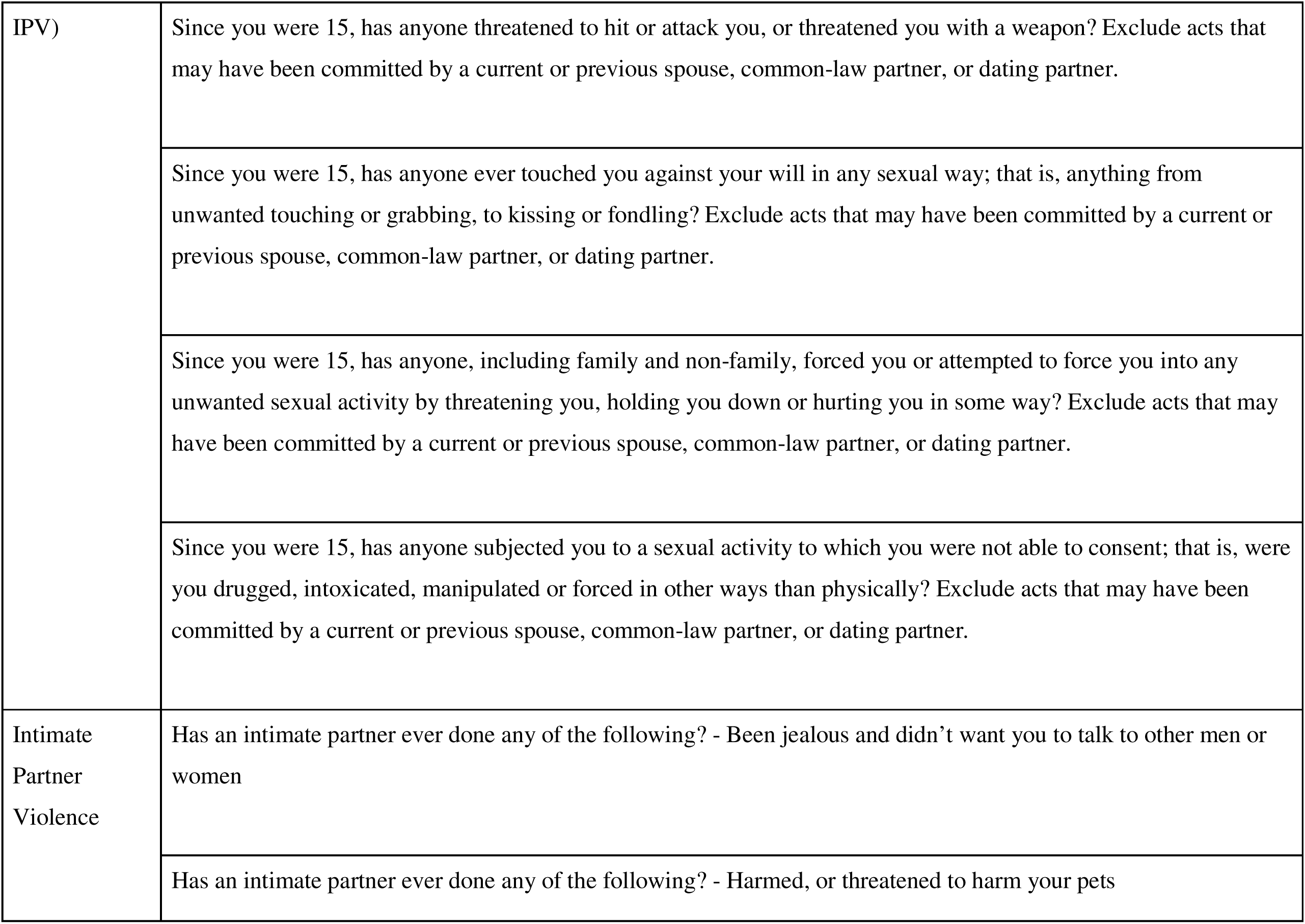
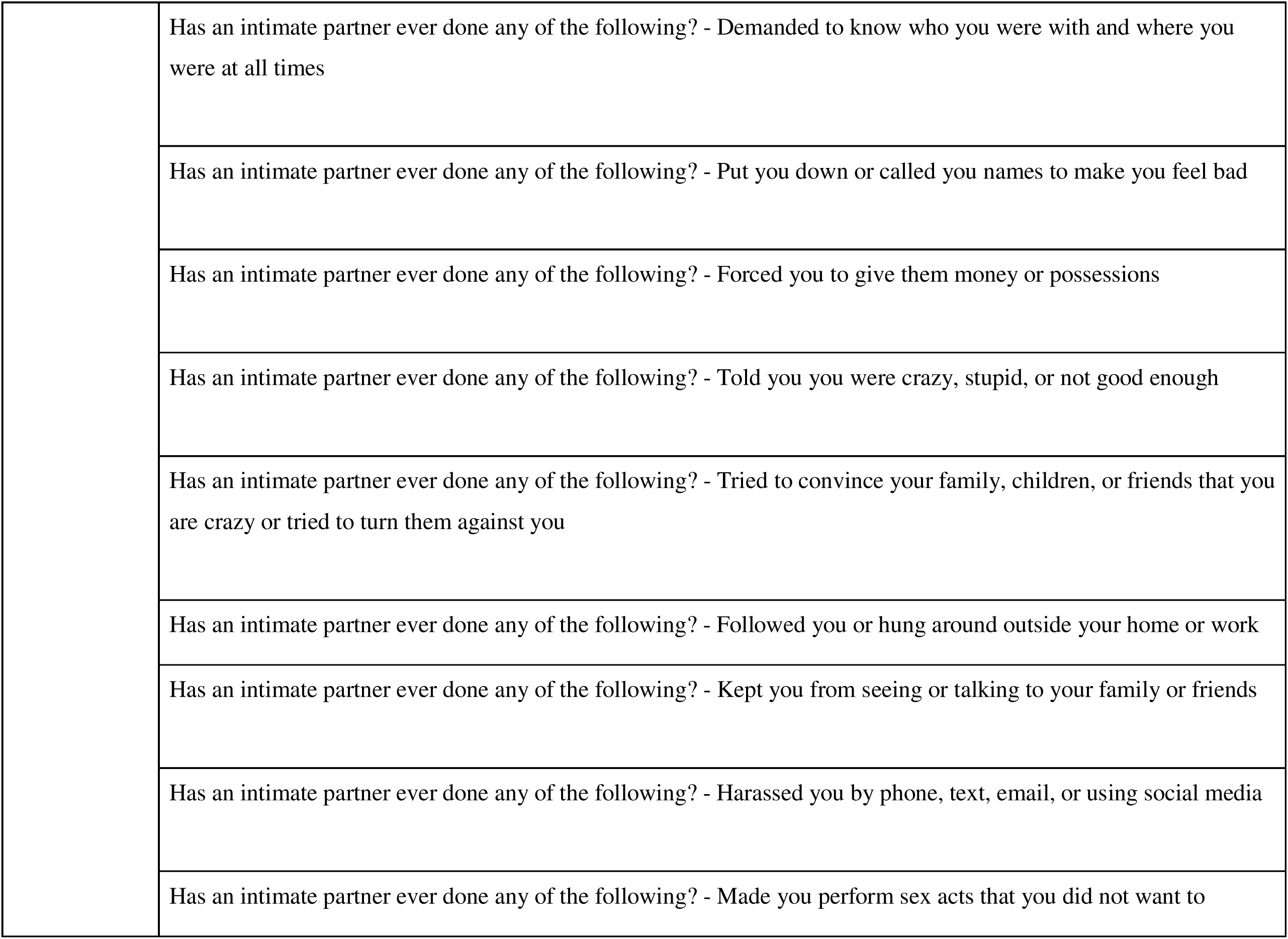
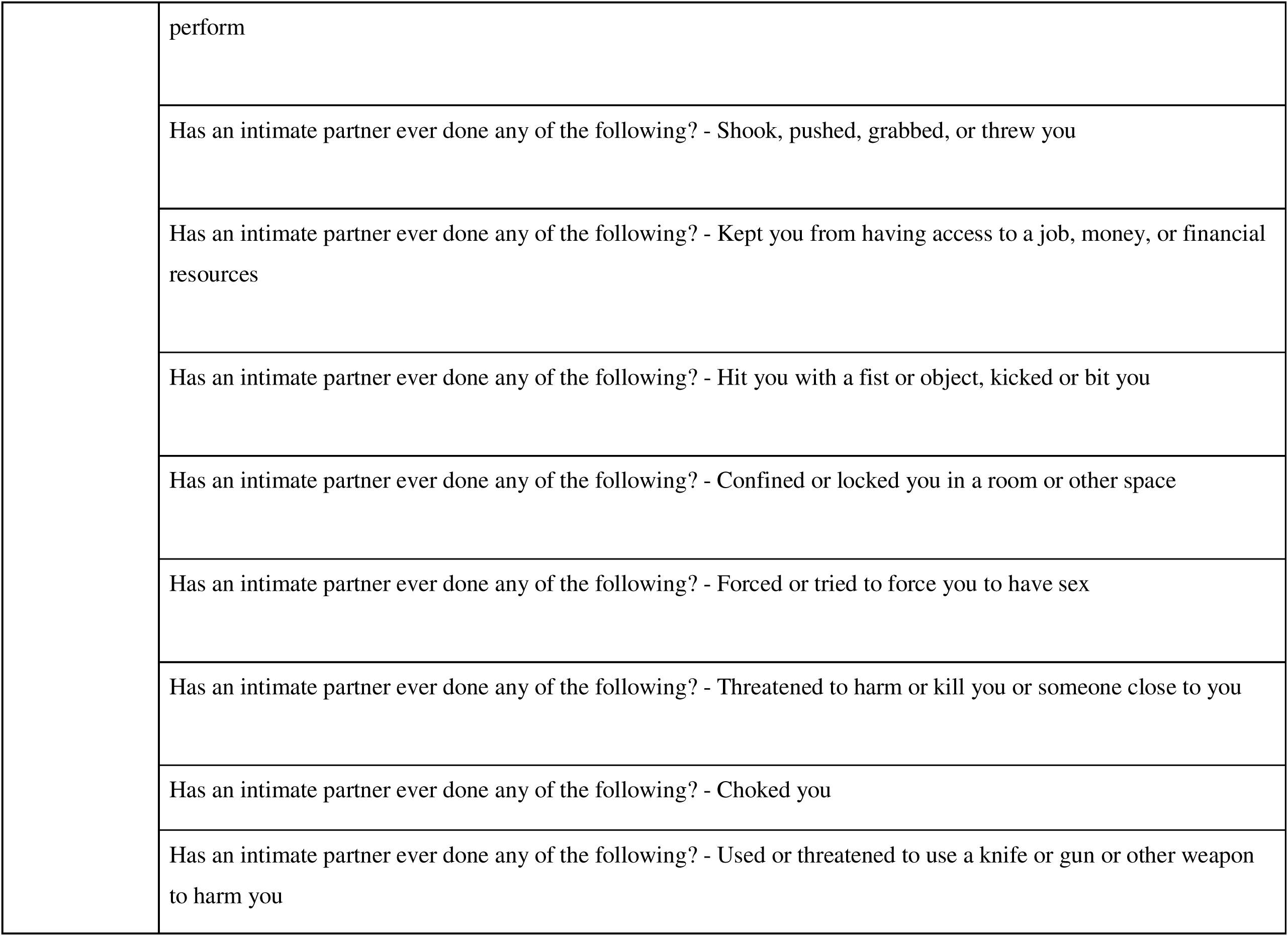
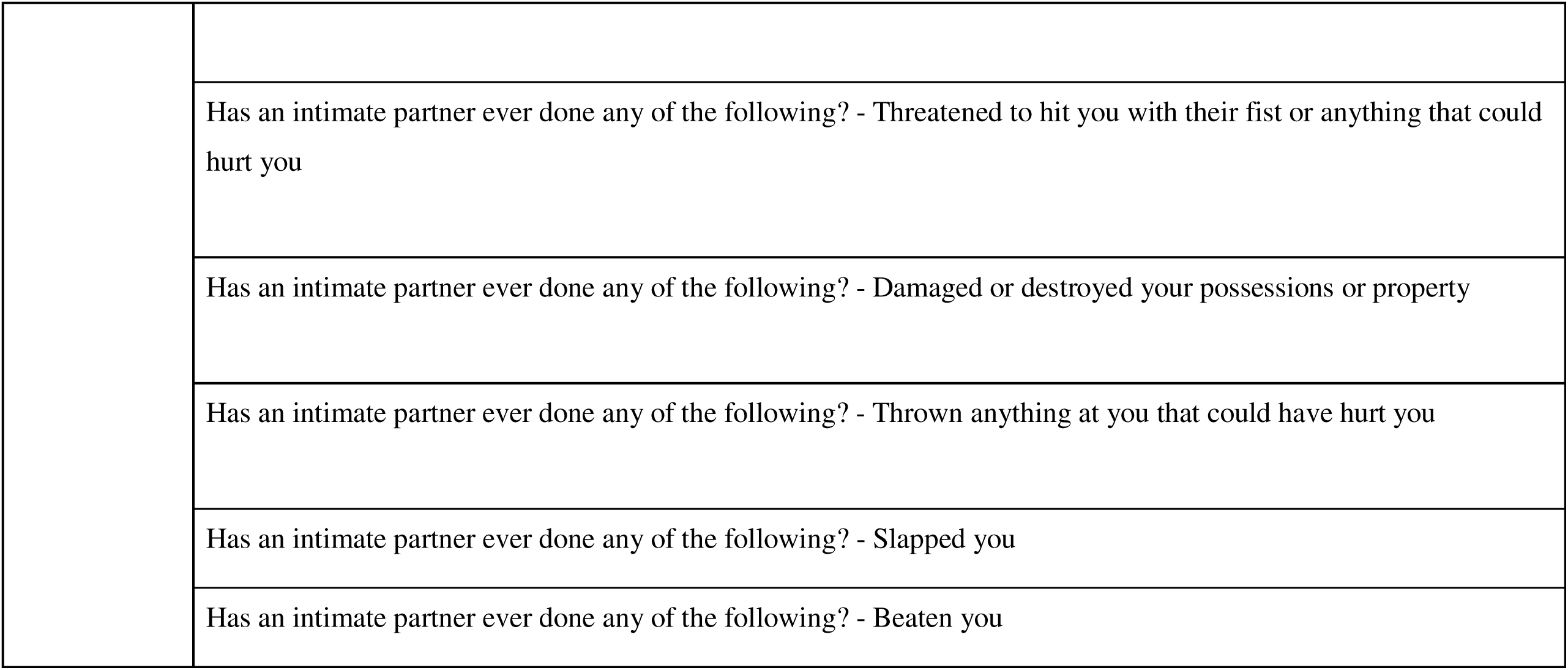
Lifetime Violence Exposure Across Childhood, Adulthood, and Intimate Partner Violence: Canadian Survey on Safety in Private and Public Spaces (SSPPS), 2018.

|  |  |
| --- | --- |
| Childhood victimization by an adult | Before age 15, how many times did any adult do any of the following to you? - Slap you on the face, head or ears, or hit you with something hard to hurt you |
|  | Before age 15, how many times did any adult do any of the following to you? - Push, grab, shove or throw something at you to hurt you |
|  | Before age 15, how many times did any adult do any of the following to you? - Kick, bite, punch, choke, burn you, or physically attack you in some way |
|  | Before age 15, how many times did any adult do any of the following to you? - Force you or attempt to force you into any unwanted sexual activity, by threatening you, holding you down or hurting you in some way |
|  | Before age 15, how many times did any adult do any of the following to you? - Touch you against your will in any sexual way, meaning anything from unwanted touching or grabbing, to kissing or fondling |
| Violent victimization as an adult (excluding | Since you were 15, have you been attacked by anyone; that is anything from being threatened, hit, slapped, pushed or grabbed, to being shot or beaten? Exclude acts that may have been committed by a current or previous spouse, common-law partner, or dating partner. |
| IPV) | Since you were 15, has anyone threatened to hit or attack you, or threatened you with a weapon? Exclude acts that may have been committed by a current or previous spouse, common-law partner, or dating partner. |
|  | Since you were 15, has anyone ever touched you against your will in any sexual way; that is, anything from unwanted touching or grabbing, to kissing or fondling? Exclude acts that may have been committed by a current or previous spouse, common-law partner, or dating partner. |
|  | Since you were 15, has anyone, including family and non-family, forced you or attempted to force you into any unwanted sexual activity by threatening you, holding you down or hurting you in some way? Exclude acts that may have been committed by a current or previous spouse, common-law partner, or dating partner. |
|  | Since you were 15, has anyone subjected you to a sexual activity to which you were not able to consent; that is, were you drugged, intoxicated, manipulated or forced in other ways than physically? Exclude acts that may have been committed by a current or previous spouse, common-law partner, or dating partner. |
| Intimate<br>Partner<br>Violence | Has an intimate partner ever done any of the following? - Been jealous and didn't want you to talk to other men or women |
|  | Has an intimate partner ever done any of the following? - Harmed, or threatened to harm your pets |
|  | Has an intimate partner ever done any of the following? - Demanded to know who you were with and where you were at all times |
|  | Has an intimate partner ever done any of the following? - Put you down or called you names to make you feel bad |
|  | Has an intimate partner ever done any of the following? - Forced you to give them money or possessions |
|  | Has an intimate partner ever done any of the following? - Told you you were crazy, stupid, or not good enough |
|  | Has an intimate partner ever done any of the following? - Tried to convince your family, children, or friends that you are crazy or tried to turn them against you |
|  | Has an intimate partner ever done any of the following? - Followed you or hung around outside your home or work |
|  | Has an intimate partner ever done any of the following? - Kept you from seeing or talking to your family or friends |
|  | Has an intimate partner ever done any of the following? - Harassed you by phone, text, email, or using social media |
|  | Has an intimate partner ever done any of the following? - Made you perform sex acts that you did not want to |
|  | perform |
|  | Has an intimate partner ever done any of the following? - Shook, pushed, grabbed, or threw you |
|  | Has an intimate partner ever done any of the following? - Kept you from having access to a job, money, or financial resources |
|  | Has an intimate partner ever done any of the following? - Hit you with a fist or object, kicked or bit you |
|  | Has an intimate partner ever done any of the following? - Confined or locked you in a room or other space |
|  | Has an intimate partner ever done any of the following? - Forced or tried to force you to have sex |
|  | Has an intimate partner ever done any of the following? - Threatened to harm or kill you or someone close to you |
|  | Has an intimate partner ever done any of the following? - Choked you |
|  | Has an intimate partner ever done any of the following? - Used or threatened to use a knife or gun or other weapon to harm you |
|  | Has an intimate partner ever done any of the following? - Threatened to hit you with their fist or anything that could hurt you |
|  | Has an intimate partner ever done any of the following? - Damaged or destroyed your possessions or property |
|  | Has an intimate partner ever done any of the following? - Thrown anything at you that could have hurt you |
|  | Has an intimate partner ever done any of the following? - Slapped you |
|  | Has an intimate partner ever done any of the following? - Beaten you |

**Supplementary Table 2:** Exposure to Violence Over Life Course by Gender.

| Supplementary Table 2: Exposure to Violence Over Life Course by Gender |  |  |  |
| --- | --- | --- | --- |
| Childhood Abuse (All Types) [CEXABU] | Women | Men | Total |
| None | 70.2% | 73.3% | 71.8% |
| Physical AND Sexual Abuse | 6.5% | 2.6% | 4.5% |
| Physical OR Sexual Abuse | 23.3% | 24.0% | 23.7% |
| Total | 100.0% | 100.0% | 100.0% |
| Childhood Sexual Abuse [CEXSEX] | **Women** | **Men** | **Total** |
| No | 87.6% | 96.1% | 91.9% |
| Yes | 12.4% | 3.9% | 8.1% |
| Total | 100.0% | 100.0% | 100.0% |
| Childhood Physical Abuse [CEXPHYS] | **Women** | **Men** | **Total** |
| No | 76.2% | 74.6% | 75.4% |
| Yes | 23.8% | 25.4% | 24.6% |
| Total | 100.0% | 100.0% | 100.0% |
| Adult Violent Victimization (Excluding IPV) [VVIC_LT] | **Women** | **Men** | **Total** |
| No | 76.2% | 74.6% | 75.4% |
| Yes | 23.8% | 25.4% | 24.6% |
| Total | 100.0% | 100.0% | 100.0% |
| Respondent experienced violent victimization (physical assault) since age 15, excluding intimate partner violence. [PA_LT] | **Women** | **Men** | **Total** |
| Yes | 76.2% | 74.6% | 75.4% |
| No | 23.8% | 25.4% | 24.6% |
| Total | 100.0% | 100.0% | 100.0% |
| Respondent experienced sexual assault since the age of 15, | **Women** | **Men** | **Total** |
| excluding intimate partner violence. [SA_LT] |  |  |  |
| Yes | 53.7% | 62.8% | 58.3% |
| No | 46.3% | 37.2% | 41.7% |
| Total | 100.0% | 100.0% | 100.0% |
| **IPV as Adult (IPV_LT)** | **Women** | **Men** | **Total** |
| No | 53.7% | 62.8% | 58.3% |
| Yes | 46.3% | 37.2% | 41.7% |
| Total | 100.0% | 100.0% | 100.0% |
| Respondent has experienced one or more forms of emotional, psychological, or financial abuse committed by an intimate partner since the age of 15. [IPVEMOLT] | **Women** | **Men** | **Total** |
| No | 53.7% | 62.8% | 58.3% |
| Yes | 46.3% | 37.2% | 41.7% |
| Total | 100.0% | 100.0% | 100.0% |
| Experienced one or more forms of physical abuse committed by an intimate partner in their lifetime (IPVPHYLT) | **Women** | **Men** | **Total** |
| No | 76.7% | 83.1% | 79.9% |
| Yes | 23.3% | 16.9% | 20.1% |
| Total | 100.0% | 100.0% | 100.0% |
| Experienced one or more forms of sexual abuse committed by an intimate partner in their lifetime (IPVSEXLT) | **Women** | **Men** | **Total** |
| No | 88.1% | 98.0% | 93.1% |
| Yes | 11.9% | 2.0% | 6.9% |
| Total | 100.0% | 100.0% | 100.0% |
| Any Violence over the Life Course (Yes to any of the above categories) [OVERALL PREVALENCE] | **Women** | **Men** | **Total** |
| No | 35.5% | 40.9% | 38.2% |
| Yes | 64.5% | 59.1% | 61.8% |
| Total | 100.0% | 100.0% | 100.0% |

**Supplementary Table 3:** Exposure to Violence Over Life Course by Past-year Economic Activity.

|  |  |  |  |
| --- | --- | --- | --- |
| Experienced Any Childhood (<15) Abuse by Adult | Did Not Work<br>Past Year | Worked Past<br>Year | Total |
| No | 68.3% | 72.4% | 71.8% |
| Physical and/or Sexual Abuse | 31.7% | 27.6% | 28.2% |
| Total | 100.0% | 100.0% | 100.0% |
| Experienced Childhood (<15) Physical Abuse by Adult | Did Not Work<br>Past Year | Worked Past<br>Year | Total |
| No | 88.2% | 92.5% | 91.9% |
| Yes | 11.8% | 7.5% | 8.1% |
| Total | 100.0% | 100.0% | 100.0% |
| Experienced Childhood (<15) Sex Abuse by Adult | Did Not Work<br>Past Year | Worked Past<br>Year | Total |
| No | 72.4% | 75.9% | 75.4% |
| Yes | 27.6% | 24.1% | 24.6% |
| Total | 100.0% | 100.0% | 100.0% |
| Violent Victimization (All Types & Excluding IPV) Since Age 15 | Did Not Work<br>Past Year | Worked Past<br>Year | Total |
| No | 60.1% | 58.5% | 58.7% |
| Yes | 40.0% | 41.5% | 41.3% |
| Total | 100.0% | 100.0% | 100.0% |
| Physical Assault (Excluding IPV) Since Age 15 | Did Not Work<br>Past Year | Worked Past<br>Year | Total |
| No | 70.0% | 66.2% | 66.7% |
| Yes | 30.0% | 33.8% | 33.3% |
| Total | 100.0% | 100.0% | 100.0% |
| Sexual Assault (Excluding IPV) Since Age 15 | Did Not Work<br>Past Year | Worked Past<br>Year | Total |
| No | 76.3% | 78.9% | 78.5% |
| Yes | 23.7% | 21.1% | 21.5% |
| Total | 100.0% | 100.0% | 100.0% |

| IPV Since 15 - All Types | Did Not Work<br>Past Year | Worked Past<br>Year | Total |
| --- | --- | --- | --- |
| No | 60.7% | 57.9% | 58.3% |
| Yes | 39.3% | 42.1% | 41.7% |
| Total | 100.0% | 100.0% | 100.0% |
| IPV Since Age 15 - Emotional/Financial Abuse | Did Not Work<br>Past Year | Worked Past<br>Year | Total |
| No | 62.3% | 59.1% | 59.6% |
| Yes | 37.7% | 40.9% | 40.4% |
| Total | 100.0% | 100.0% | 100.0% |
| IPV Since Age 15 - Sexual Abuse | Did Not Work<br>Past Year | Worked Past<br>Year | Total |
| No | 91.4% | 93.4% | 93.1% |
| Yes | 8.7% | 6.6% | 6.9% |
| Total | 100.0% | 100.0% | 100.0% |

| IPV Since Age 15 - Physical Abuse | Did Not Work<br>Past Year | Worked Past<br>Year | Total |
| --- | --- | --- | --- |
| No | 78.5% | 80.2% | 79.9% |
| Yes | 21.5% | 19.8% | 20.1% |
| Total | 100.0% | 100.0% | 100.0% |
| Lifetime Violence Exposure | Did Not Work<br>Past Year | Worked Past<br>Year | Total |
| No | 39.2% | 38.0% | 38.2% |
| Yes | 60.8% | 62.0% | 61.8% |
| Total | 100.0% | 100.0% | 100.0% |

Note: All comparisons are made with unexposed men.

**Supplementary Table 4:**
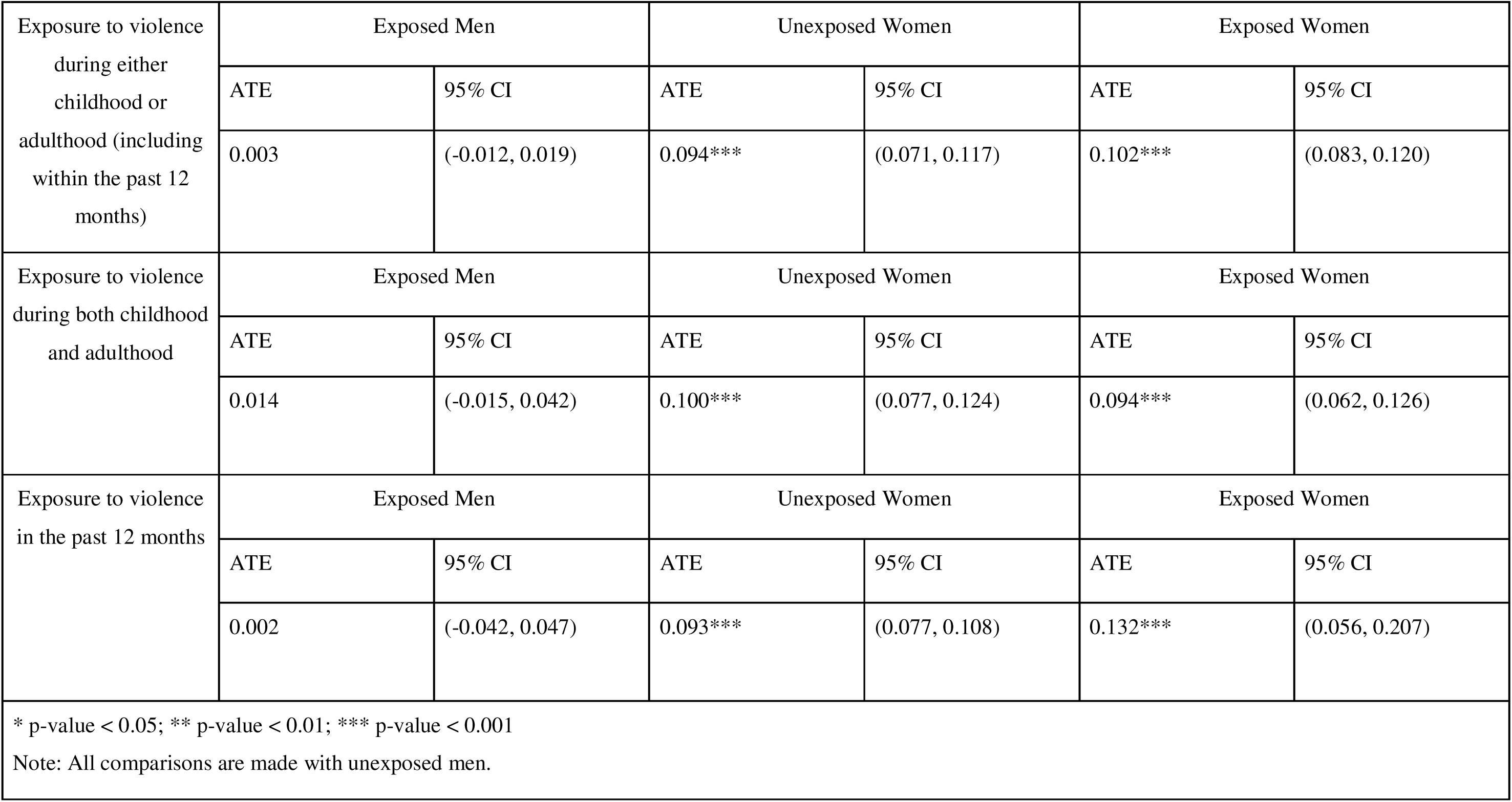
IPWRA Results for Log Odds of Past-year Economic Inactivity Stratified by Type of Exposure.

